# *HFE* p.C282Y (rs1800562) allele frequencies in 34 population/control cohorts in Iberia

**DOI:** 10.64898/2025.12.19.25342681

**Authors:** James C. Barton, J. Clayborn Barton, Ronald T. Acton

## Abstract

**Background:** *HFE* p.C282Y (c.845G>A; rs1800562) is a common missense mutation in persons of European ancestry, but we found no comprehensive tabulation of p.C282Y allele frequencies in Iberia.

**Methods:** We performed computerized and manual searches to identify evaluable reports of p.C282Y alleles in population/control cohorts ≥50 subjects in Iberia. We tabulated numbers of subjects, nominal geographic sites of cohort recruitment, cohort characteristics, corresponding latitudes and longitudes, and p.C282Y allele frequencies [95% confidence intervals]. We computed the aggregate p.C282Y allele frequencies of mainland Spain and mainland Portugal and compared the aggregate frequencies using the Chi-square test (two-tailed). Using combined mainland Spain and mainland Portugal data, we computed the aggregate p.C282Y allele frequency in Iberia.

**Results:** We identified 25 cohorts in mainland Spain (12,297 subjects; 11 of the 15 autonomous communities) and nine cohorts in mainland Portugal (1,024 subjects; each of the five administrative regions). Cohorts were recruited in this region: latitude 43.4619 – 37.2299° N; longitude -9.1366 – 2.1899° W. The range of p.C282Y allele frequencies in the 34 cohorts was 0.0000 to 0.0517. The aggregate p.C282Y allele frequency in mainland Spain was 0.0291 (716/24,594) [0.0271, 0.0313] and that in mainland Portugal was 0.0303 (62/2048) [0.0237, 0.0386] (p = 0.8343). The aggregate p.C282Y allele frequency in Iberia was 0.0292 (778/26,642) [0.0272, 0.0313].

**Conclusions:** We conclude that the aggregate *HFE* p.C282Y allele frequencies in mainland Spain and mainland Portugal do not differ significantly. The aggregate p.C282Y allele frequency of 34 population/control cohorts (13,321 subjects, 16 geographic regions) in Iberia is 0.0292.

## Introduction

*HFE*, the homeostatic iron regulator (chromosome 6p22.2) [1,2], encodes the non-classical class I major histocompatibility complex protein HFE, an upstream modulator of the central iron-regulatory hormone hepcidin (*HAMP*, chromosome 19q13.12) [3]. *HFE* p.C282Y (c.845G>A; rs1800562) is a common missense mutation in persons of European ancestry [4,5]. p.C282Y homozygosity is associated with the predominant subtype of hemochromatosis [2,6].

Iberia, also known as the Iberian Peninsula, is a landmass of 583,544 km^2^ (225,308 square miles) in southwestern Europe, which is separated from the rest of Europe by the Pyrenees Mountains [7]. The area of Iberia is predominantly that of mainland Spain (84.5%) and mainland Portugal (15.3%) [7]. The remaining area in the northeast comprises the microstate Andorra and a small part of the French department of Pyrénées-Orientales, and in the south, Gibraltar, a British Overseas Territory [7]. A comprehensive tabulation of the *HFE* p.C282Y allele frequencies of population/control cohorts in Iberia could be used to enhance knowledge of the geographic distribution of p.C282Y.

The aims of this study were 1) to identify all published reports of *HFE* p.C282Y allele frequencies in population/control cohorts in Iberia, 2) to tabulate the sizes, geographic locations, characteristics, and p.C282Y allele frequencies of the evaluable cohorts, and 3) to compute the aggregate p.C282Y allele frequency in Iberia. We discuss our findings and compare the aggregate p.C282Y allele frequency in Iberia with previously published p.C282Y allele frequencies in other regions of Europe.

## Methods

### Ethics statement

This study was performed according to the principles of the Declaration of Helsinki [8]. This study did not require institutional review board approval because the reports we identified were published previously and are publicly available, the report data contain no personally identifiable information, and the tabulation of the data does not permit re-identification of individuals [9].

### Informed consent statement

Informed consent was not obtained because this study was based entirely on the discovery, review, tabulation, and analyses of published data that are publicly available and cannot be linked to individuals [9].

### Definition of population/control cohort

We defined a population cohort as a group of research subjects who share a common characteristic(s) and are used in a study to represent the broader population. We defined a control cohort as a group of individuals in a study who do not have the condition or outcome of interest but who are otherwise similar to the individuals in the main study group and are presumed to represent the broader population. In this study, we defined population and control cohorts to be equivalent.

### Literature searches for population/control cohorts

We performed computerized searches (using an internet browser with or without https://pubmed.ncbi.nlm.nih.gov/) to identify reports of *HFE* p.C282Y allele frequencies in population/control cohorts in Iberia. We used *HFE* p.C282Y, rs1800562, and *HFE* mutation as search terms in combination with 1) names of countries, autonomous communities, administrative regions, and major cities, 2) disease-related terms including hemochromatosis, iron overload, cirrhosis, diabetes, arthritis, hepatitis, osteoporosis, porphyria cutanea tarda, leukemia, lymphoma, myeloma, cancer, and carcinoma, and 3) the terms *HFE* p.H63D (rs1799945) and *HFE* p.S65C (rs1800730). We manually searched the references cited in reports identified by computerized searches.

### Definition of evaluable population/control cohorts

We defined evaluable cohorts as ones in which the corresponding reports included all of the following data: 1) 50 or more population/control subjects [10]; 2) the nominal geographic site of subject recruitment (or location of the primary investigator’s institution, as appropriate); 3) the characteristics of the subjects; and 4) determinable numbers of *HFE* p.C282Y and total alleles.

### Population/control cohorts excluded

We did not tabulate data from reports that described the following: 1) Roma people and other Iberian residents who were not regarded as Iberian natives by the corresponding investigators; 2) no geographic location of cohort recruitment other than country; 3) *HFE* p.C282Y allele frequencies estimated using population prevalences of p.C282Y homozygotes; and 4) the publication of a previously reported population/control cohort. We excluded cohorts from the Balearic and Canary Islands (Spain), the Azores and Madeira (Portugal), and the autonomous cities of Ceuta and Melilla in Northern Africa (Spain) because these geographic regions are not in Iberia [7].

### Data tabulation

We tabulated these data of the evaluable population/control cohorts: 1) the numbers of subjects; the nominal geographic sites of the recruitment of subjects (or the locations of primary investigator institutions, as appropriate); 3) the characteristics of the cohorts; 4) the latitudes and longitudes in decimal degrees (four decimal places) [11] of the geographic sites of recruitment; and 5) the corresponding *HFE* p.C282Y allele frequencies.

### Statistics

All data are displayed herein and in the corresponding literature citations. We computed the *HFE* p.C282Y allele frequency for each cohort as the quotient of (number of p.C282Y alleles) by (number of subjects x 2), expressed to four decimal places [95% confidence interval]. We computed the aggregate p.C282Y allele frequencies for mainland Spain and mainland Portugal in a similar manner and compared them using the Chi-square test (two-tailed). We defined allele frequencies from mainland Spain and mainland Portugal as representative of Iberia and likewise computed the aggregate p.C282Y allele frequency for Iberia. We used Excel^®^ 2000 (Microsoft Corp., Redmond, WA, USA) and GraphPad Prism 8^®^ (2018; GraphPad Software, San Diego, CA, USA). We defined p < 0.05 as significant.

## Results

### Mainland Spain

We identified 25 evaluable population/control cohorts (12,297 subjects) from these 11 autonomous communities of mainland Spain: Aragon, Asturias, Basque Country, Cantabria, Castile-La Mancha, Catalonia, Extremadura, Galicia, Madrid, Murcia, and Valencia (Table 1). The range of subjects per cohort was 50 to 5,370 (Table 1). Cohorts were recruited in this region: latitude 43.4619 – 37.9924° N; longitude -8.5459 – 2.1771° W (Table 1). The range of *HFE* p.C282Y allele frequencies was 0.0068 (Murcia/Murcia) to 0.0517 (Basque Country/Guipuzcoa) (Table 1).

**Table 1.** *HFE* p.C282Y allele frequencies in 12,082 population/control subjects in mainland Spain.

| Subjects, n | Autonomous community/ province or city | Latitude° N | Longitude° W | p.C282Y/ total alleles | p.C282Y allele frequency [95% confidence interval] | Author (Year) [reference] |
| --- | --- | --- | --- | --- | --- | --- |
| 215 | Aragon/Zaragoza<sup>1</sup> | 41.6521 | -0.8809 | 430 | 0.0372 [0.0230, 0.0596] | Solano-Barca (2009) [12] |
| 159 | Asturias/Oviedo<sup>2</sup> | 43.3619 | -5.8484 | 11/318 | 0.0346 [0.0194, 0.0609] | Lauret (2002) [13] |
| 51 | Basque Country<sup>3</sup> | 42.9912 | -2.5543 | 2/102 | 0.0196 [0.0054, 0.0687] | Baiget (1998) [14] |
| 116 | Basque Country/Guipuzcoa<sup>4</sup> | 43.1445 | -2.2038 | 12/232 | 0.0517 [0.0298, 0.0882] | de Juan (2001) [15]<sup>5</sup> |
| 281 | Basque Country/Barakaldo<sup>6</sup> | 43.2955 | -2.9901 | 12/562 | 0.0214 [0.0123, 0.0370] | de Buruaga (2023) [17] |
| 213 | Cantabria/Santander<sup>7</sup> | 43.4619 | -3.8100 | 19/426 | 0.0446 [0.0287, 0.0686] | Fábrega (2004) [18] |
| 150 | Castile-La Mancha/Toledo<sup>8</sup> | 39.8559 | -4.0243 | 9/300 | 0.0300 [0.0159, 0.0560] | de Diego (2004) [19] |
| 50 | Catalonia<sup>9</sup> | 41.8523 | 1.5745 | 3/100 | 0.0300 [0.0103, 0.0845] | Merryweather-Clarke (1997) [4] |
| 348 | Catalonia<sup>10</sup> | 41.8523 | 1.5745 | 21/696 | 0.0302 [0.0198, 0.0457] | Berez (2005) [20] |
| 108 | Catalonia/Barcelona<sup>11</sup> | 41.3826 | 2.1771 | 8/216 | 0.0370 [0.0189, 0.0713] | Baiget (1998) [14]<sup>12</sup> |
| 512 | Catalonia/Barcelona<sup>13</sup> | 41.3826 | 2.1771 | 31/1024 | 0.0303 [0.0214, 0.0427] | Sánchez (1998) [22] |
| 5370 | Catalonia/Barcelona<sup>14</sup> | 41.3826 | 2.1771 | 339/10740 | 0.0316 [0.0285, 0.0351] | Sánchez (2003) [23] |
| 1043 | Catalonia/Barcelona<sup>15</sup> | 41.3826 | 2.1771 | 61/2086 | 0.0292 [0.0228, 0.0373] | Altes (2004) [24] |
| 126 | Catalonia/Barcelona<sup>16</sup> | 41.3826 | 2.1771 | 7/252 | 0.0278 [0.0135, 0.0562] | Toll (2006) [25] |

<sup>1</sup> normolipemic subjects
<sup>2</sup> healthy bone marrow donors
<sup>3</sup> not otherwise specified
<sup>4</sup> provincial blood donors with Basque surnames
<sup>5</sup> Castiella et al. (2010) published similar data [16].

|  |  |  |  |  |  |  |
| --- | --- | --- | --- | --- | --- | --- |
| 812 | Catalonia/Tarragona<sup>17</sup> | 41.1172 | 1.2546 | 50/1624 | 0.0308 [0.0234, 0.0404] | Aranda (2007) [26]<sup>18</sup> |
| 179 | Extremadura/Badajoz<sup>19</sup> | 38.8782 | -6.9701 | 16/358 | 0.0447 [0.0277, 0.0714] | Rodríguez-López (2012) [28] |
| 125 | Extremadura/Cáceres<sup>20</sup> | 39.5152 | -6.4506 | 5/250 | 0.0200 [0.0086, 0.0460] | Alvarez (2001) [29] |
| 50 | Galicia/Santiago de Compostela<sup>21</sup> | 42.8804 | -8.5459 | 5/100 | 0.0500 [0.0215, 0.1118] | Soto (2000) [30] |
| 50 | Madrid/Madrid<sup>22</sup> | 40.4583 | -3.7727 | 2/100 | 0.0200 [0.0055, 0.0700] | de Salamanca<sup>23</sup> (1999) [32] |
| 174 | Madrid/Madrid<sup>24</sup> | 40.4583 | -3.7727 | 8/348 | 0.0230 [0.0117, 0.0447] | Moreno (1999) [33] |
| 150 | Madrid/Madrid<sup>25</sup> | 40.4583 | -3.7727 | 9/300 | 0.0300 [0.0159, 0.0560] | Gonzalez-Hevilla (2005) [34] |
| 551 | Madrid/Madrid<sup>26</sup> | 40.4583 | -3.7727 | 25/1102 | 0.0227 [0.0154, 0.0333] | Ropero-Gradilla (2005) [35] |
| 1000 | Madrid/Madrid<sup>27</sup> | 40.4583 | -3.7727 | 33/2000 | 0.0165 [0.0118, 0.0231] | Ropero (2006) [36] |
| 370 | Murcia/Murcia<sup>28</sup> | 37.9924 | -1.1305 | 5/740 | 0.0068 [0.0029, 0.0158] | Muro (2007) [37] |
| 94 | Valencia/Valencia<sup>29</sup> | 39.4697 | -0.3763 | 7/188 | 0.0372 [0.0181, 0.0748] | Monzó (2017) [38] |

<sup>17</sup> randomly selected from the electoral rolls of three municipalities in Tarragona
<sup>18</sup> Aranda et al. (2010) published similar data [27].
<sup>19</sup> unrelated gender-matched healthy individuals
<sup>20</sup> voluntary unrelated blood donors
<sup>21</sup> random samples representative of the Galician population

We did not discover reports of evaluable population/control cohorts from these four autonomous communities of north-central and south-central mainland Spain: Andalusia, Castile and León, La Rioja, and Navarre.

### Mainland Portugal

We identified nine evaluable population/control cohorts (1,024 subjects) from the five administrative regions of mainland Portugal: Alentejo, Algarve, Centro, Lisbon-Tagus Valley, and Norte (Table 2). The range of subjects per cohort was 52 to 146 (Table 2). Cohorts were recruited in this region: latitude 41.4563 – 37.2299° N; longitude -9.1366 – -7.4817° W (Table 2). The range of *HFE* p.C282Y allele frequencies was 0.000 to 0.0581 (Table 2).

**Table 2.**
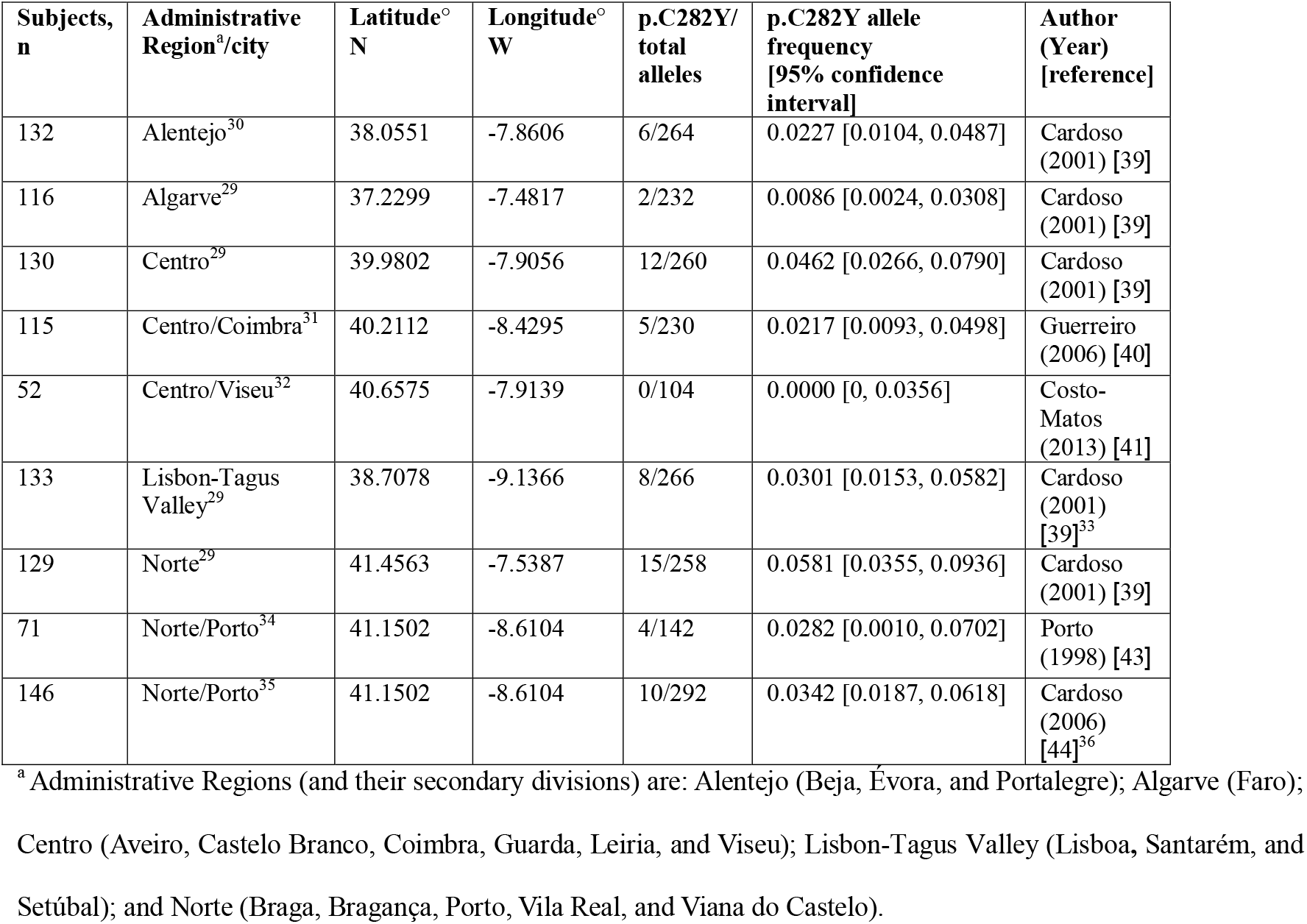
*HFE* p.C282Y allele frequencies in 1,024 population/control subjects in mainland Portugal.

### Other regions of Iberia

We did not discover reports of evaluable population/control cohorts from Andorra, Pyrénées-Orientales, or Gibraltar.

### Aggregate HFE p.C282Y allele frequencies

The aggregate p.C282Y allele frequency in mainland Spain was 0.0291 (716/24,594) [0.0271, 0.0313] (Table 1). The aggregate p.C282Y allele frequency in mainland Portugal was 0.0303 (62/2048) [0.0237, 0.0386] (Table 2). These frequencies did not differ significantly (p = 0.8343). The aggregate p.C282Y allele frequency in Iberia was 0.0292 (778/26,642) [0.0272, 0.0313].

## Discussion

Novel findings of this tabulation of *HFE* p.C282Y allele frequencies in Iberia are 1) the large number of evaluable population/control cohorts we discovered and 2) the wide range of p.C282Y allele frequencies across geographic regions. The present aggregate p.C282Y allele frequency percentage in Iberia (2.9) is lower than the p.C282Y allele frequency percentages reported from Ireland (9.7, 10.0), Wales (8.2), Brittany (7.7), Scotland (7.7), England (6.0, 7.7), and Norway (6.4, 6.6), and higher than the p.C282Y frequency percentages reported from Greece (1.0, 1.4) and Italy (0.5, 1.7) [4,5]. The present tabulation of p.C282Y allele frequencies in 34 Iberian cohorts (13,321 subjects, 16 geographic regions) could be used to enhance knowledge of the distribution of p.C282Y in Iberia and Europe.

Uncertainties of this study include the possibilities that: 1) we overlooked one or more published reports of evaluable population/control cohorts; 2) the same subjects were included in more than one cohort or report; and 3) some cohorts were not representative of their respective geographic regions due to the site(s) or method(s) of subject recruitment or to inadequate cohort size, especially in geographic regions with low *HFE* p.C282Y allele frequencies. A limitation of this study is that we discovered no reports of evaluable population/control cohorts from five autonomous communities of mainland Spain or from Andorra, Pyrénées-Orientales, or Gibraltar. Analyzing the relationships of the present p.C282Y allele frequencies with their geographic locations in Iberia or with regional prevalences of hemochromatosis associated with p.C282Y homozygosity was beyond the scope of this study.

## Conclusions

We conclude that the aggregate *HFE* p.C282Y allele frequencies in mainland Spain and mainland Portugal do not differ significantly. The aggregate p.C282Y allele frequency of 34 population/control cohorts (13,316 subjects, 16 geographic regions) in Iberia is 0.0292.

## Data Availability

All data are displayed herein and in the corresponding literature citations.

## Authors’ contributions

Each author contributed substantively to this study. JaCB conceived this study and its methodology, curated data, evaluated reports from the literature, performed analyses, and drafted the manuscript. JClB curated data, evaluated reports from the literature, performed analyses, and drafted the manuscript. RTA curated data, evaluated reports from the literature, contributed to study methodology, performed analyses, and drafted the manuscript. Each author approved the manuscript in its final form.

## Data Sharing Statement

All data used in the present study are either displayed herein or in the corresponding literature citations.

## Footnotes

1 normolipemic subjects

2 healthy bone marrow donors

3 not otherwise specified

4 provincial blood donors with Basque surnames

5 Castiella et al. (2010) published similar data [16].

6 controls with gout (n = 128), osteoarthritis (n = 102), and rheumatoid arthritis (n = 70) without chondrocalcinosis, calcium pyrophosphate crystals in synovial fluid, hemochromatosis, or primary hyperparathyroidism

7 provincial blood donors

8 randomly selected healthy subjects without iron overload

9 anthropological community-based survey

10 randomly selected subjects from population registries in two villages

11 voluntary blood donors

12 Fernndez-Real et al. (1999) [21] and Gimferrer et al. (1999) [14] published similar data.

13 voluntary blood donors and paternity testing clients

14 voluntary blood donors

15 randomly selected newborn screening cards

16 randomly selected laboratory subjects (76 healthy + 50 with chronic hepatitis C without a history of porphyria cutanea tarda or hemochromatosis)

17 randomly selected from the electoral rolls of three municipalities in Tarragona

18 Aranda et al. (2010) published similar data [27].

19 unrelated gender-matched healthy individuals

20 voluntary unrelated blood donors

21 random samples representative of the Galician population

22 unrelated healthy subjects

23 Ramirez et al. (2009) reported similar data [31].

24 randomly selected subjects

25 normal subjects

26 cord blood leukocytes

27 neonates

28 unrelated white Spanish blood donors

29 random healthy subjects

30 from a stratified random sample of anonymous dried blood spot samples from newborns

31 healthy controls free of clinical signs or symptoms of neurodegeneration

32 healthy controls (during elective cholecystectomy)

33 Machado et al. (2009) published similar data [42].

34 unrelated apparently healthy subjects

35 apparently healthy control population of women

36 Bettencourt et al. (2011) published similar data [45].

